# Surveillance genome sequencing reveal multiple SARS-CoV-2 variants circulating in the central Texas, USA with a predominance of Delta variant and review of vaccine breakthrough cases

**DOI:** 10.1101/2021.08.06.21261727

**Authors:** Manohar B. Mutnal, Shelby Johnson, Nada Mohamed, Rasha Abddelgader, Linden Morales, Marcus Volz, Kimberly Walker, Alejandro C. Arroliga, Arundhati Rao

## Abstract

As surges in the COVID-19 pandemic have continued worldwide, SARS-CoV-2 has mutated, spawning several new variants, and impacting, to various degrees, transmission, disease severity, diagnostics, therapeutics, and natural and vaccine-induced immunity. Baylor Scott & White Health has implemented, along with laboratory diagnosis, SARS-CoV-2 sequencing to identify variants in its geographical service area. We analyzed virus sequencing results of specimens collected across Central Texas and found dramatic changes in variant distribution in the first half of 2021. The alpha variant (B 1.1.7) became predominant at week 13 and continued dominance until week 25. A growth rate of 1.20 (R^2^ = 0.92) for the first 15 weeks was noted and this growth gradually declined to -0.55 (R^2^ = 0.99) for the final 13 weeks. Currently, B.1.1.7 is being displaced with B.1.617.2 at 0.58 growth rate (R^2^ = 0.97). We also investigated vaccine breakthrough cases within our healthcare system and present clinical data on 28 symptomatic patients.

## Introduction

As of July 2021, severe acute respiratory syndrome coronavirus 2 (SARS-CoV-2) has resulted in over 4.1 million deaths worldwide, more than 600,000 in the United States of America, and more than 51,000 in the state of Texas alone.(1) As the pandemic continues to surge in countries that have not yet received meaningful supplies of vaccines and in communities that remain resistant to vaccination, genetic variants are emerging that have different characteristics in terms of transmissibility, the severity of disease they cause, and, potentially, their susceptibility to the treatments and vaccines developed based on the wild-type virus.(2) Of particular concern are emerging variants that include mutations in the S gene, coding for the viral spike protein, which has been the primary target of the most effective vaccines and passive antibody treatments authorized and deployed against COVID-19.(3) Variants with mutations in the S gene are potential threats to their continued efficacy.(4, 5) These concerns have provided an impetus to increase testing and sequencing of viral DNA in infected persons in order to understand the transmissibility, virulence, and ability of variants to evade current vaccines (6).

RNA viruses exist as a swarm or “quasispecies” of genome sequences around a core consensus sequence (7). Under conditions of selection, such as those imposed by neutralizing antibodies or drugs, variants of the swarm can escape genetically and become resistant. To date, 4150 mutations have been identified in the S gene of SARS-CoV-2 isolated from humans ([CoV-GLUE, 2021](https://www.biorxiv.org/content/10.1101/2020.11.06.372037v2.full#ref-7); [GISAID, 2021](https://www.biorxiv.org/content/10.1101/2020.11.06.372037v2.full#ref-12)). These mutations give rise to 1,246 amino acid changes including 187 substitutions in the receptor binding domain (RBD) (8). The abundance of many variants in the human population suggests they are not accompanied by a fitness loss. Multiple mechanisms likely account for the emergence of such substitutions including host adaptation, immune selection during natural infection, and possibly reinfection of individuals with incomplete or waning immunity.

Most clinical laboratories performing diagnostic testing for SARS-CoV-2 are not equipped to detect genetic variants unless additional resources are made available for genomic sequencing of suspected cases and false negatives caused by mutations in the targeted genes. The lack of funding for technology, clinical utility and regulatory oversight may have contributed to non-availability of sequencing methods in the clinical laboratories. Epidemiological investigations are not part of the diagnostic laboratory routine workflow; however, the widespread nature of SARS-CoV-2 pandemic necessitates clinical laboratories be prepared – through funding, technology acquisition, appropriate staffing, and technical competency - to perform genetic sequencing to aid in the control measures.

To assist public health officials involved in tracking and tracing the SARS-CoV-2 variants in the Central Texas community, the Baylor Scott & White Health (BSWH) molecular microbiology laboratory participated in genotyping of previously confirmed SARS-CoV-2 specimens. The reports generated were not released to patients’ electronic health records but were provided to public health officials for necessary interventions when a variant of concern (VOC) or variant of interest (VOI) was identified. Similarly, vaccine breakthrough cases were identified and reported according to Centers for Disease Control and Prevention (CDC) definitions.(9)

In this report, we present evolution of SARS-CoV-2 lineages in the Central Texas area, and their association with vaccine breakthrough cases, along with clinical history of those breakthrough cases. In large, randomized-controlled trials, each vaccine was found to be safe and efficacious in preventing symptomatic, laboratory-confirmed COVID-19 (10) (11). In this report, we present evolution of SARS-CoV-2 lineages in the limited geography of Central Texas and their association with vaccine breakthrough along with clinical history of vaccinated patients exposed to variants.

## Material and Methods

This study was reviewed and approved by the Baylor Scott and White Research Institute Institutional Review board (IRB # 021-144) with a waiver of the requirement of informed consent for the use of residual specimens for sequencing and epidemiological studies.

### Specimen Selection for sequencing

Since the beginning of the pandemic our laboratory has archived all the positive specimens confirmed by one of the nucleic acid amplification methods that were deployed for diagnosis of SARS-CoV-2 infections. Since the implementation of sequencing, both retrospective and prospective specimens were selected for variant identification in our community. BSWH molecular pathology laboratory has conducted approximately 400,000 tests and SARS-CoV-2 and reported 10.8% overall positivity rate for the Central Texas geography. These tests include both symptomatic and surveillance specimens since the beginning of the pandemic, March 2020. While surveillance testing resulted in 1.8% and symptomatic 15.8% positivity rates (Data accessed on 27^th^, May 2021). Genome sequencing was introduced to detect SARS-CoV-2 variants from the beginning of the year, 2021.

Specimens (nasopharyngeal swabs) positive for SARS-CoV-2 by emergency use authorization (EUA)-cleared nucleic acid amplification methods, performed at Department of Pathology & Laboratory Medicine, Baylor Scott & White Hospital, Temple, TX from 1/1/2021 to 07/07/2021, were identified for sequencing. Specimens for sequencing were selected based on Ct <30 on a polymerase chain reaction (PCR) method, or RLU >1000 on a transcription mediated amplification (TMA) method. We randomly selected 1% of the positive specimens for sequencing when positivity rates were higher than 5% and all positive specimens when positivity rates were <5%, to identify the variants in circulation.

### Vaccine breakthrough cases

An electronic query was set up in the electronic health record to identify patients with a COVID-19 vaccination history. If patients were tested multiple times post vaccination they were distinctly counted, and the first positive was counted for statistical purposes.

### Library Preparation and Sequence Data Analysis

Nucleic acid was purified from each specimen and subjected to reverse transcription, next-generation sequencing library preparation, sequencing, and data analysis according to the manufacturer’s recommendation using two different platforms;

### Sequencing on Ion Torrent (ThermoFisher) S5 system

Extracted RNA was subject to reverse transcription using the SuperScript VILO cDNA Synthesis Kit Chef (ThermoFisher Scientific, Waltham, MA). Library preparation and subsequent templates were performed using the Ampliseq SARS-CoV-2 Research Panel, DL8 kit, and the Ion 510 & Ion 520 & Ion 530 Kit on the Ion Chef. Sequencing was performed on the Ion Torrent S5. Data were manually uploaded to the NextClade bioinformatics pipeline for analysis.

### Sequencing on Illumina COVIDSeq System

Extracted RNA was subject to reverse transcription and library preparation using the Illumina COVIDSeq Test protocol and reagents (Illumina, Inc, San Diego, CA). Libraries were pooled per manufacturer instruction and sequenced on the Illumina NextSeq. Data was analyzed using the Illumina BaseSpace application/bioinformatics pipeline, DRAGEN COVID Lineage v3.5.1.

### Data collection

Epidemiological, demographic, clinical and laboratory data were extracted from electronic health records and the laboratory information system.

### Statistical Analysis

Descriptive analysis was performed to present demographics factors (age, gender, and ethnicity), clinical presentation, clinical outcome, and molecular studies on SARS-CoV-2 specimens. Growth rate statistical analyses were performed in SAS 9.4. All graphs were created in R version 3.5.1. To calculate growth curves, the Verhulst growth model, also known as the logistic growth model, was was used to model population growth with constraints. The formula for the Verhulst growth model is:

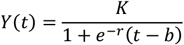

Where: *Y*(*t*) = The population size at a given time, *K* = The upper limit of the population size (in our case, 1), *r* = the rate of maximum growth, *b* = the time at which the quantity is half its maximum value, *t* = time

## Results

### Evolution of Central Texas variants and rapid displacement of B.1.1.7 with B.1.617.2

Since the beginning of the pandemic our laboratory has archived all the positive specimens confirmed by one of the nucleic acid amplification methods that were deployed for diagnosis of SARS-CoV-2 infections. Since the implementation of sequencing, both retrospective and prospective specimens were selected for variant identification in our community. BSWH molecular Microbiology laboratory has conducted approximately 400,000 tests and SARS-CoV-2 and reported 10.8% overall positivity rate for the Central Texas geography. These tests include both symptomatic and surveillance specimens since the beginning of the pandemic, March 2020. While surveillance testing resulted in 1.8% and symptomatic 15.8% positivity rates (Data accessed on 27^th^, May 2021). Genome sequencing was introduced to detect SARS-CoV-2 variants from the beginning of the year, 2021. We randomly selected 1% of the positive specimens for sequencing during initial weeks when positivity rates was higher than 5% and once the overall positivity rate was below 5% all positive specimens were in the pipeline for sequencing to identify the variants in circulation.

Figure 1 shows the distribution of variants in circulation by week of collection during the study period. Data presented in the Figure 1 do not reflect every sample sequenced as there is a process lag.

**1.**
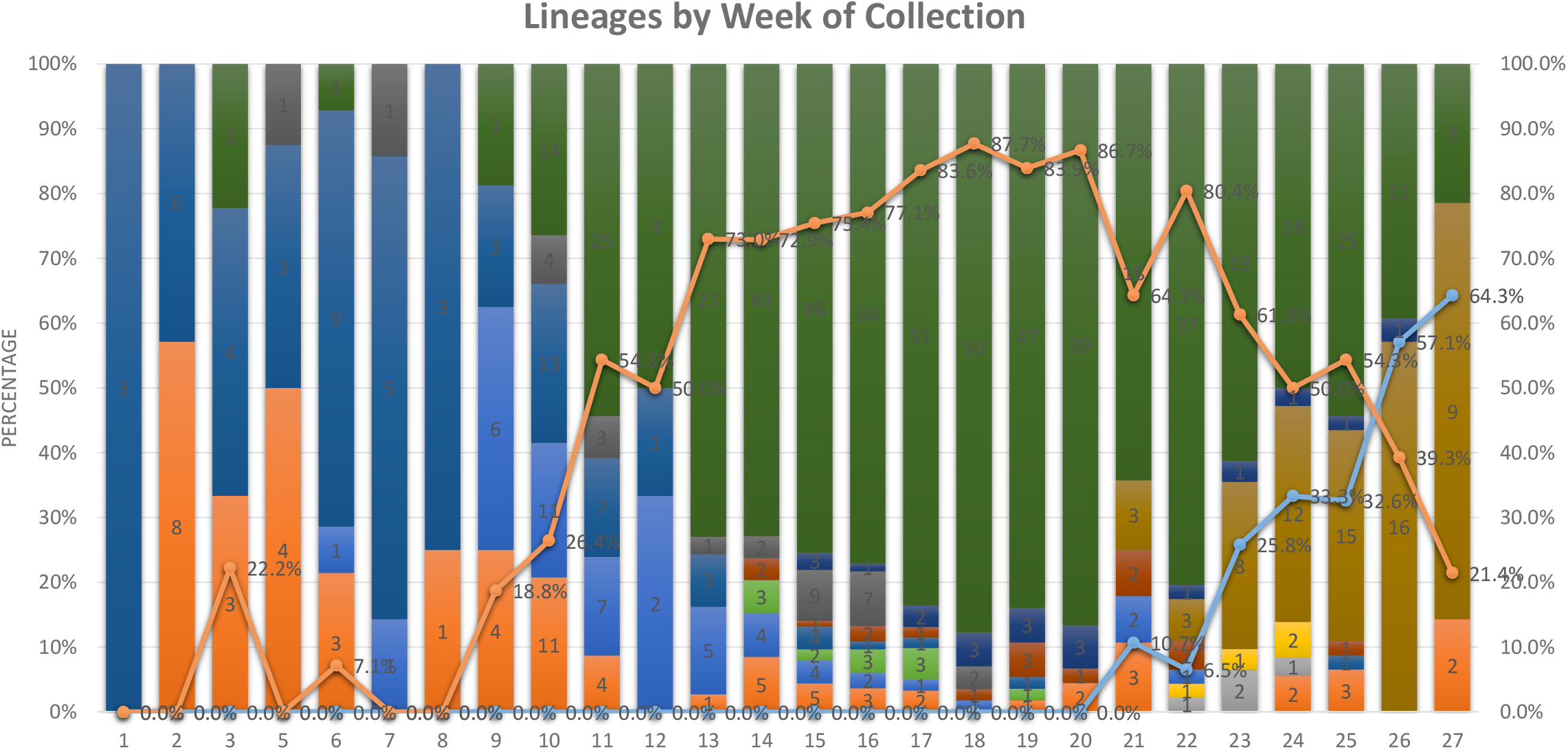
SARS-CoV-2 variant distribution. Nucleic Acid Amplification confirmed SARS-CoV-2 specimens were frozen until sequenced. Data presented in the bar graph shows variant distribution for the respective weeks and the trend lines for Alpha and Delta variants.

The alpha variant (B 1.1.7) became predominant variant at week 13 and continued dominance until week 25 (Figure 2A). A growth rate of 1.20 (R^2^ = 0.92) for the first 15 weeks was noted and this growth gradually declined to -0.55 (R^2^ = 0.99) for the final 13 weeks. Currently, B.1.1.7 is being displaced with the delta variant (B.1.617.2) at 0.58 growth rate (R^2^ = 0.97) (Figure 2B)

**2.**
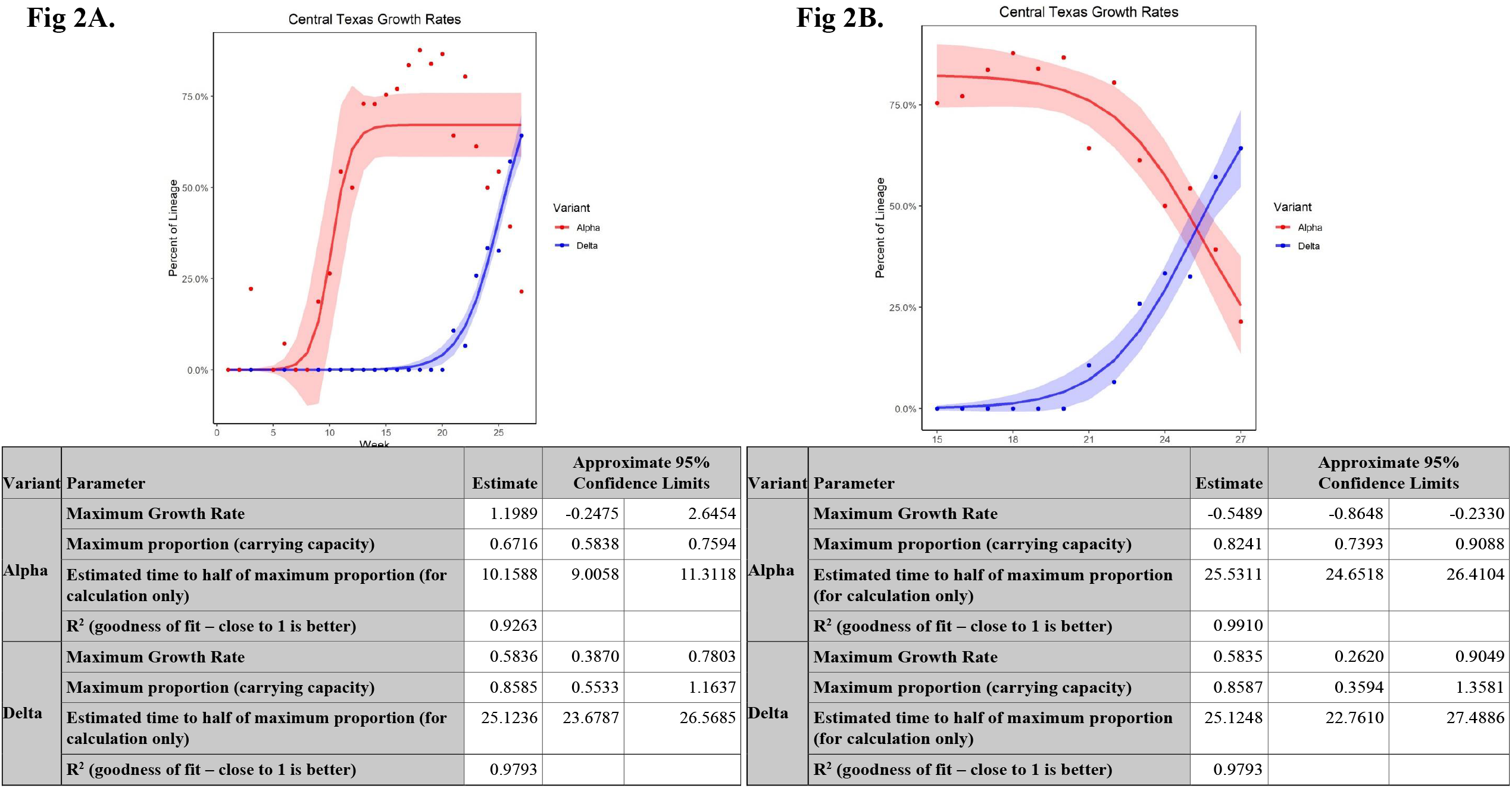
Growth curves for Alpha and Delta variants. Growth rate statistical analyses were performed in SAS 9.4. All graphs were created in R version 3.5.1. To calculate growth curves, the Verhulst growth model, also known as the logistic growth model was used to model population growth with constraints. Data presented in Fig 2A shows growth curves for both Alpha and Delta variants from the beginning of the year 2021 and Fig 2B rates for both variants for final 13 weeks.

### Vaccine breakthrough studies

BSWH tested 96,357 patients for SARS-CoV2 by nucleic acid amplification testing (NAAT) between 01/01/2021 and 05/31/2021. Patients tested multiple times were distinctly counted once. In the event a patient was tested multiple times with both detected and not detected results, only the first detected result was counted for capturing the vaccine breakthrough on the earliest day of detection.

Among the 96,357 patients, 3925 (4%) were fully vaccinated with the BNT162b2, mRNA-1273, or Janssen COVID-19 Vaccine, and were at least 14 days post the final dose. An additional 3164 (3.2%) patients were partially vaccinated either with BNT162b2 or mRNA-1273. Patients were considered partially vaccinated if their second dose (or the single dose for Janssen COVID-19) was <14 days before the specimen collection being investigated.

According to the established criteria (https://www.cdc.gov/vaccines/covid-19/health-departments/breakthrough-cases.html), we detected 72 (1.8%) vaccine breakthrough cases out of 3925 fully vaccinated individuals. Of these 72 cases, 33 (45.8%) specimens had <30 Ct value or >1000 RLU on the TMA method and so were suitable for sequencing. In the partially vaccinated patients, 207 vaccine breakthrough cases were detected for a 6.54% positivity rate, while among the 90,898 unvaccinated individuals, 10,503 (11.5%) were positive for SARS-CoV-2 infection.

Data presented in Figure 3 show the weekly distribution of vaccine breakthrough cases and the variants involved. The vaccine breakthrough investigation was begun at week 10. The alpha variant (B 1.1.7) caused 16 (22.2%) out 72 vaccine breakthrough cases, while the delta variant (B.1.617.2) contributed 11 (15.2%) vaccine breakthrough cases, all during the final 4 weeks of the study period (week 24-27), as this variant gained more prevalence.

**Figure 3.**
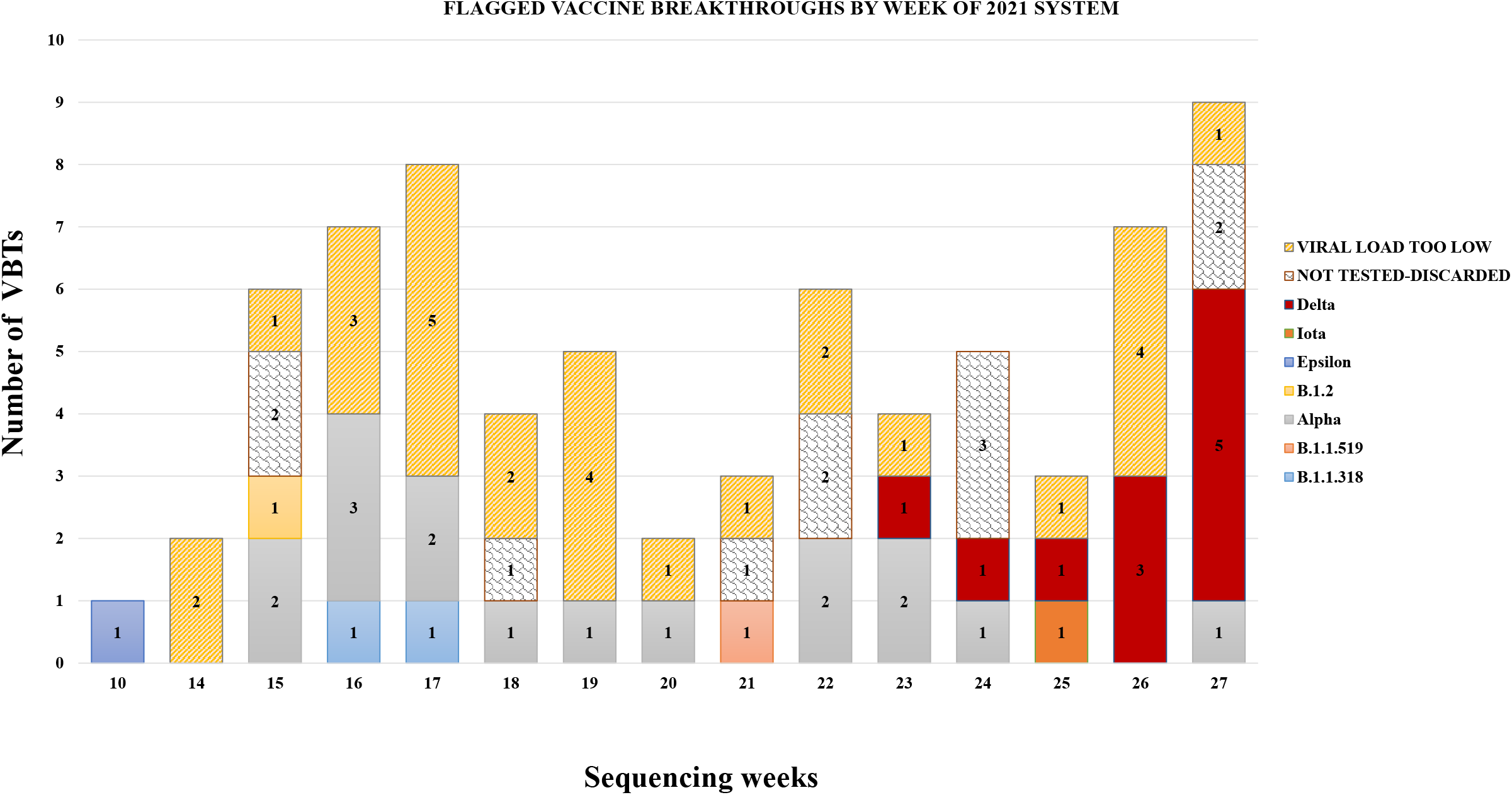
Vaccine breakthrough cases. Data presented in the bar graph shows vaccine breakthrough cases for respective sequencing weeks.

### Clinical presentation of vaccine breakthrough cases

We performed a review of electronic health records of patients who were fully vaccinated and experienced vaccine breakthrough (Table 1). Among the 33 vaccine breakthrough cases that were sequenced, 28 were included in the VBT study as their clinical data existed in our EHR. Of the 28 VBT cases reviewed, 17 (60%) were females, and the mean age of these 28 patients was 54.9 (20 – 88 years).

**Table I.** Demographic, comorbidity, symptoms, other social and past medical history of BSWH vaccine breakthrough cases.

| Baseline characteristics (total n=28) |  |  |  |
| --- | --- | --- | --- |
| Demographics |  |  |  |
| Gender %(n) | Male 39% (11) |  | Female 60% (17) |
| Age (years) | Mean 54.9 |  | Range 20-88 |
| Comorbid conditions |  |  |  |
| Type 2 Diabetes Mellitus | 28.6% (8) |  |  |
| Asthma %(n) | 17.9% (5) |  |  |
| COPD %(n) | 7.1% (2) |  |  |
| CHF %(n) | 3.6% (1) |  |  |
| HTN %(n) | 39.3% (11) |  |  |
| Current Smokers | 10.7 (3) |  |  |
| Former Smokers | 35.7% (10) |  |  |
| Subjective symptoms |  |  |  |
|  | BNT162b2 | Janssen COVID-19 Vaccine | mRNA-1273 |
|  | TOTAL 17 | TOTAL 11 | TOTAL 0 |
| Asymptomatic 28.6% (8) | 41.2% (7) | 9.1% (1) |  |
| Fever/Chills 25% (7) | 17.6% (3) | 36.4% (4) |  |
| Headache 28.6% (8) | 23.5% (4) | 36.4% (4) |  |
| Rhinorrhea 21.4% (6) | 5.9% (1) | 45.5% (5) |  |
| Cough 46.4% (13) | 23.5% (4) | 81.8% (9) |  |
| Otalgia/pressure 3.6% (1) | 0% (0) | 9.1% (1) |  |
| Odynophagia 17.9% (5) | 17.6% (3) | 18.2% (2) |  |
| SOB/Severe dyspnea 14.3% (4) | 5.9% (1) | 27.3% (3) |  |
| Diarrhea 7.1% (2) | 5.9% (1) | 9.1% (1) |  |
| Loss of taste and smell 14.3% (4) | 5.9% (1) | 27.3% (3) |  |

Most of the vaccine breakthrough patients (96%) had comorbid conditions, hypertension being the most common (39.3%). Other common comorbid conditions were determined and diabetes mellitus (28.6%), asthma (17.9%), and chronic obstructive pulmonary disease (7.1%). Approximately 45% of the patients reported being either active (10%) or former smokers (35%).

Table 1 shows the symptoms associated with breakthrough under each vaccine brand in our sample. However, direct comparisons between brands cannot be made with our data, given both the small numbers and the complexities related to the different dosing schedules and different rates of use in the population, and conclusions about the relative effectiveness of the different brands overall or against specific variants should not be inferred from these results. Two (2.1%) vaccine breakthrough cases were hospitalized with shortness of breath.

## Discussion

In this study, we examined the SARS-CoV-2 variant distribution in the Central Texas region in the first half of 2021, finding first growing dominance by the alpha (B.1.1.7) variant, with a maximum growth rate of 1.20 in the first 15 weeks, followed by rapid displacement by the delta (B1.617.2) variant at a 0.58 growth rate. We also identified 207 vaccine breakthrough cases among the 3164 partially vaccinated individuals, and 72 among the 3925 fully vaccinated individuals for whom we had testing samples, including 28 for whom the variant with which they were infected was able to be determined. Similar to the overall pattern of various distribution among COVID cases in this study population, the sequenced vaccine breakthrough cases showed a predominance of the alpha (B.1.1.7) variant being rapidly replaced by the delta (B.1.617.2) variant in the last few weeks of the study period.

Our findings regarding the distribution of circulating variants in Central Texas are similar to those observed for the United States as a whole. The alpha (B.1.1.7) variant became the predominant variant across the United States by April 2021, followed by rapid displacement by the delta (B.1.617.2) variant, which became the dominant strain early in July 2021. (12, 13) By July 3, 2021, the delta (B.1.617.2) variant accounted for an estimated 61.7% of COVID-19 cases nationally, (13) which is similar to the 64% we observed in the final week of our study period (ending on 7/7/2021). Such rapid, widespread changes in variant distribution underscore the critical need for robust and timely genomic surveillance (14). In this study, we showed SARS-CoV-2 variant distribution in the Central Texas region and rapid displacement of current B.1.1.7 (Alpha) variant with B1.617.2 (Delta) variant. The distribution of circulating SARS-CoV-2 variants in the United States changed rapidly during the period of January 2020 – May 2021. The expansion of the Alpha variant to become the predominant variant in all U.S. regions in the beginning of the year 2021, and the more recent emergence of the Delta variant in all regions, underscore the critical need for robust and timely genomic surveillance (14). A growth rate of 1.20 for the first 15 weeks was noted for Alpha variant and this growth gradually declined to - 0.55 for the final 13 weeks. Currently, B.1.1.7 is being displaced with B.1.617.2 at 0.58 growth rate. Our data collected from Central TX reflects the national trend that Delta is rapidly becoming prevalent variant (12).

Questions raised with each new variant spreading include whether the variant will “escape” the protection conferred by either natural immunity following infection with a different strain of the virus or acquired immunity from one of the authorized vaccines, all of which were developed based on the version of Sars-CoV-2 originally detected in Wuhan, China. Of particular interest in the United States, currently, is whether the predominance of the delta (B.1.617.2) variant will cause an increased rate of vaccine breakthrough cases. In the spring of 2021, the alpha (B.1.1.7) variant plateaued at relatively low level in the many U.S. regions in the context of increasing vaccination while mask mandates and social distancing measures implemented by local and state governments, as well as many private businesses, remained in place. Texas, which ended government-ordered mask mandates and other pandemic-related restrictions in March 2021,(15) has seen a rapid rise in COVID-19 cases and hospitalizations again in July 2021,(16, 17) concomitant with the rising dominance of the delta (B.1.617.2) variant. Our results examining the variant distribution in vaccine breakthrough cases show that both the alpha (B.1.1.7) and delta (B.1.617.2) variants have caused some small number of breakthrough cases, with predominance patterns appearing to mirror those of the broader cases of COVID-19. The number of samples suitable for sequencing in our study is too small, and the portion of our study period in which the delta (B.1.617.2) variant held dominance too short, to make any comparisons regarding the risk for vaccine breakthrough these two variants carry.

Almost all (96%) of the fully vaccinated patients who experienced vaccine breakthrough in our study had comorbid chronic conditions. Little evidence is currently available regarding risk factors for vaccine breakthrough infection, but our results are consistent with such evidence as is available. One study conducted using data from the Veterans Health Administration reported that increasing age, multiple comorbid conditions, and non-black race were associated with confirmed breakthrough infection.(18) A systematic review of studies reporting serious adverse effects, SARS-CoV-2 infection and deaths after COVID-19 vaccination, identified comorbidities, Variants of Concern, and casual attitude towards COVID Appropriate Behaviors among the most important factors contributing to risk for vaccine breakthrough infection.(19)

We also examined the symptoms the 28 fully vaccinated patients with breakthrough infections experienced. Cough was most commonly reported (46.4%), but four (14.3%) had shortness of breath and 2 (7%) required hospitalization. In terms of serious illness requiring hospitalization, these results are similar to those reported by the CDC from 46 U.S. states and territories as for January 1 to April 30, 2021, in which 10% of patients with breakthrough infections required hospitalization.(20) Likewise, similar percentages of breakthrough cases were asymptomatic in our sample (28.6%) and in the national data reported by the CDC (27%).(20)

Our study design was not intended to deduce vaccine effectiveness against any particular variant, but the small numbers of vaccine breakthrough infections we identified add to the growing evidence that the greatest remaining risks for COVID-19 in the United States are almost entirely isolated to the unvaccinated. Equally important is the substantially higher positivity rate we observed in partially vaccinated individuals. It is to be hoped that continued accumulation of such evidence will encourage the 40% of eligible Texans who remain entirely unvaccinated, and the 9% who have not completed their vaccination schedule,(21) to take this important step in protecting themselves, their families, and their communities. Continued monitoring will, of course, be needed, particularly given the current overall rise in case numbers in Texas, and the recent shift in dominance from the alpha (B.1.1.7) to the delta (B.1.617.2) variant. A second limitation that must be kept in mind when interpreting the results reported here is that, while clinical manifestation data for the 28 vaccine breakthrough cases for which sequencing was possible are presented by vaccine brand they cannot support comparisons of effectiveness between the brands, as there was unequal distribution of vaccine brands in the Central Texas community.

In conclusion, our results show that, in early 2021, Central Texas experienced a rapid growth of the alpha (B.1.1.7) variant, which then maintained dominance for several weeks, but towards the end of the study period (June/July 2021) was itself rapidly displaced by the delta (B.617.2) variant. Examination of vaccine breakthrough infections showed small numbers overall and with each of these variants, with the distribution of the variants among breakthrough cases following a similar pattern to the distribution in the overall COVID-19 cases in our study population. Continued monitoring is needed, both to assess the longer-term impact of the delta (B.1.617.2) variant’s dominance on risk for vaccine breakthrough infections, and to identify any new variants, or variants altering the distribution in circulating strains, that might impact risks for disease and/or effectiveness of vaccines and therapeutics.

Lastly, the findings in this report are subject to at least two limitations. First, the number of reported COVID-19 vaccine breakthrough cases is likely a substantial undercount of all SARS-CoV-2 infections among fully vaccinated persons. This study relied on electronic health records of patients within our healthcare system and data might not be complete or representative. Many persons with vaccine breakthrough infections, especially those who are asymptomatic or who experience mild illness, might not seek testing. Second, SARS-CoV-2 sequence data are available for only a small proportion of the reported cases.

## Data Availability

All data is available for review and is hosted on the secure network drive of the organization.

## Acknowledgment

Authors would like to thank Kendall Hammonds for help with statistical analysis and Briget Da Graca for editorial help.

## References

1. COVID-19 Map - Johns Hopkins Coronavirus Resource Center.

2. Sanyaolu A, Okorie C, Marinkovic A, Haider N, Abbasi AF, Jaferi U, Prakash S, Balendra V. 2021. The emerging SARS-CoV-2 variants of concern. Ther Adv Infect Dis 8:20499361211024372.

3. Aleem A, Akbar Samad AB, Slenker AK. 2021. Emerging Variants of SARS-CoV-2 And Novel Therapeutics Against Coronavirus (COVID-19)StatPearls. StatPearls Publishing, Treasure Island (FL).

4. Wang R, Chen J, Gao K, Wei G-W. 2021. Vaccine-escape and fast-growing mutations in the United Kingdom, the United States, Singapore, Spain, India, and other COVID-19-devastated countries. Genomics 113:2158–2170.

5. Effectiveness of Covid-19 Vaccines against the B.1.617.2 (Delta) Variant | NEJM.

6. Hacisuleyman E, Hale C, Saito Y, Blachere NE, Bergh M, Conlon EG, Schaefer-Babajew DJ, DaSilva J, Muecksch F, Gaebler C, Lifton R, Nussenzweig MC, Hatziioannou T, Bieniasz PD, Darnell RB. 2021. Vaccine Breakthrough Infections with SARS-CoV-2 Variants. N Engl J Med 384:2212–2218.

7. Dolan PT, Whitfield ZJ, Andino R. 2018. Mapping the Evolutionary Potential of RNA Viruses. Cell Host Microbe 23:435–446.

8. Liu Z, VanBlargan LA, Bloyet L-M, Rothlauf PW, Chen RE, Stumpf S, Zhao H, Errico JM, Theel ES, Liebeskind MJ, Alford B, Buchser WJ, Ellebedy AH, Fremont DH, Diamond MS, Whelan SPJ. 2021. Landscape analysis of escape variants identifies SARS-CoV-2 spike mutations that attenuate monoclonal and serum antibody neutralization. bioRxiv 2020.11.06.372037.

9. 2021. COVID-19 Breakthrough Case Investigations and Reporting | CDC.

10. Polack FP, Thomas SJ, Kitchin N, Absalon J, Gurtman A, Lockhart S, Perez JL, Pérez Marc G, Moreira ED, Zerbini C, Bailey R, Swanson KA, Roychoudhury S, Koury K, Li P, Kalina WV, Cooper D, Frenck RW, Hammitt LL, Türeci Ö, Nell H, Schaefer A, Ünal S, Tresnan DB, Mather S, Dormitzer PR, Şahin U, Jansen KU, Gruber WC, C4591001 Clinical Trial Group. 2020. Safety and Efficacy of the BNT162b2 mRNA Covid-19 Vaccine. N Engl J Med 383:2603–2615.

11. Baden LR, El Sahly HM, Essink B, Kotloff K, Frey S, Novak R, Diemert D, Spector SA, Rouphael N, Creech CB, McGettigan J, Khetan S, Segall N, Solis J, Brosz A, Fierro C, Schwartz H, Neuzil K, Corey L, Gilbert P, Janes H, Follmann D, Marovich M, Mascola J, Polakowski L, Ledgerwood J, Graham BS, Bennett H, Pajon R, Knightly C, Leav B, Deng W, Zhou H, Han S, Ivarsson M, Miller J, Zaks T, COVE Study Group. 2021. Efficacy and Safety of the mRNA-1273 SARS-CoV-2 Vaccine. N Engl J Med 384:403–416.

12. Bolze A, Cirulli ET, Luo S, White S, Cassens T, Jacobs S, Nguyen J, Ramirez JM, Sandoval E, Wang X, Wong D, Becker D, Laurent M, Lu JT, Isaksson M, Washington NL, Lee W. 2021. Rapid displacement of SARS-CoV-2 variant B.1.1.7 by B.1.617.2 and P.1 in the United States. medRxiv 2021.06.20.21259195.

13. CDC. 2020. COVID Data Tracker. Centers for Disease Control and Prevention.

14. Paul P, France AM, Aoki Y, Batra D, Biggerstaff M, Dugan V, Galloway S, Hall AJ, Johansson MA, Kondor RJ, Halpin AL, Lee B, Lee JS, Limbago B, MacNeil A, MacCannell D, Paden CR, Queen K, Reese HE, Retchless AC, Slayton RB, Steele M, Tong S, Walters MS, Wentworth DE, Silk BJ. 2021. Genomic Surveillance for SARS-CoV-2 Variants Circulating in the United States, December 2020-May 2021. MMWR Morb Mortal Wkly Rep 70:846–850.

15. Executive Order GA-34. Texas Department of State Health Services.

16. COVID-19 In Texas (Dashboard).

17. COVID-19 in Texas - Texas Tests and Hospitals (Dashboard).

18. Butt AA, Khan T, Yan P, Shaikh OS, Omer SB, Mayr F. 2021. Rate and risk factors for breakthrough SARS-CoV-2 infection after vaccination. J Infect https://doi.org/10.1016/j.jinf.2021.05.021.

19. Jain VK, Iyengar KP, Ish P. 2021. Elucidating causes of COVID-19 infection and related deaths after vaccination. Diabetes Metab Syndr 15:102212.

20. CDCMMWR. 2021. COVID-19 Vaccine Breakthrough Infections Reported to CDC — United States, January 1–April 30, 2021. MMWR Morb Mortal Wkly Rep 70.

21. Workbook: COVID-19 Vaccine in Texas (Dashboard).

